# Differential age-associated brain atrophy and white matter changes among homeless and precariously housed individuals compared to the general population

**DOI:** 10.1101/2021.11.03.21265661

**Authors:** Jacob L. Stubbs, Andrea A. Jones, Daniel Wolfman, Ryan C. Y. Chan, Alexandra T. Vertinsky, Manraj K. Heran, Wayne Su, Donna J. Lang, Thalia S. Field, Kristina M. Gicas, Melissa L. Woodward, Allen E. Thornton, Alasdair M. Barr, Olga Leonova, G. William MacEwan, Alexander Rauscher, William G. Honer, William J. Panenka

**Affiliations:** Department of Psychiatry, University of British Columbia, Vancouver, Canada; British Columbia Mental Health and Substance Use Services Research Institute, Vancouver, Canada; Division of Neurology, University of British Columbia, Vancouver, Canada; Department of Radiology, University of British Columbia, Vancouver, Canada; Department of Psychology, York University, Toronto, Canada; Department of Psychology, Simon Fraser University, Vancouver, Canada; Department of Anesthesiology, Pharmacology, and Therapeutics, University of British Columbia, Vancouver, Canada; Department of Pediatrics, University of British Columbia, Vancouver, Canada; British Columbia Neuropsychiatry Program, Vancouver, Canada

## Abstract

**Importance:** Homeless or precariously housed individuals live with poor health and experience premature mortality compared to the general population. With an increasing average age among this demographic, syndromes associated with neurogenerative disease are also increasing. Quantitative MRI measures may help define the roles of age and risk factors for poor brain health among these individuals.

**Objective:** To evaluate whether MRI measures of brain structure are differentially associated with age and selected risk factors among individuals who are homeless or precariously housed compared to a general population sample.

**Design, setting, and participants:** Cross sectional comparison of baseline data from 312 community-based, precariously housed participants with a publicly available dataset of 382 participants recruited from the general population.

**Exposure(s):** The primary exposure was housing status (precariously housed *vs* general population). Risk factors in the precariously housed sample included mental illness, substance dependence, intravenous drug use, HIV, and history of traumatic brain injury.

**Main outcome(s) and measure(s):** The main outcomes were MRI measures of whole-brain tissue-to-intracranial volume ratio, fractional anisotropy, and mean diffusivity. Multiple linear regression and piecewise regression were used to evaluate differences in associations between MRI measures and age between the samples, and to explore associations with risk factors in the precariously housed sample.

**Results:** Compared to the general population sample, older age in the precariously housed sample was associated with more whole-brain atrophy (*β*=-0.20, p=0.0029), lower whole-brain FA (*β*=-0.32, *p*<0.0001), and higher whole-brain MD (*β*=0.69, *p*<0.0001). Several MRI measures had non-linear associations with age, with further adverse changes after age 35-40 in the precariously housed sample. Frontal and temporal cortical thickness, corpus callosum volume, and diffusivity in the association tracts, corpus callosum, and thalamic radiations were the regions of interest most differentially affected. History of traumatic brain injury, stimulant dependence, and heroin dependence were associated with more atrophy or alterations in white matter diffusivity in the precariously housed sample.

**Conclusions and relevance:** Older age is associated with adverse MRI measures of brain structure among homeless and precariously housed individuals compared to the general population. Education, improvements in care provision and policy may help to reduce the health disparities experienced by these individuals.

## INTRODUCTION

Individuals who are homeless experience disproportionately high morbidity and mortality compared to the general population,^1^ and individuals who are precariously housed face similar mortality as those who are homeless.^2^ Previous studies report considerably higher rates of communicable diseases,^3^ mental illness,^4^ and substance misuse, as well as high rates of unintentional injury such as falls, assault, and traumatic brain injury.^5,6^ Previous studies also report higher rates of non-communicable diseases, cognitive impairment, and age-related conditions, which some have taken as evidence for accelerated aging.^5^ Indeed, homeless individuals over the age of 50 have a high prevalence of “geriatric syndromes”, comparable with general population samples more than 20 years older.^7,8^

There is preliminary evidence for differential brain changes in homeless and precariously housed samples, with a higher-than-expected prevalence of stroke and nearly one-third having pathological findings visible on MRI.^9,10^ Only one study to date has looked at quantitative MRI in homeless individuals compared to a control group and found that homeless individuals had smaller thalamic and brainstem volumes compared to controls, and that history of traumatic brain injury was associated with lower volumes of various brain structures compared to those without traumatic brain injury.^11^ However, this study was limited in sample size (*n*_homeless_ = 25, *n*_control_ = 26) and reported only basic between-group differences in volume across a few regions of interest. To date, it is unknown whether MRI measures of brain structure differ between homeless or precariously housed individuals compared to the general population, or whether there is a differential association with age that could indicate accelerated degenerative processes. If there are differential associations between MRI measures of brain structure and age, there are potential implications for education, public health policy, and health delivery, especially given the aging homeless population.^12^

In this study we evaluated whether age is differentially associated with MRI measures of brain structure in a well characterized sample of individuals who are precariously housed (*n* = 312) compared to a publicly available sample of individuals from the general population (*n* = 398). We hypothesized that older age would be associated with greater whole-brain atrophy and differential white matter diffusivity among precariously housed individuals compared to the general population. We characterized differential associations with age across regions of interest and also explored whether risk factors that are overrepresented in homeless or precariously housed samples, such as mental illness diagnoses, substance dependence, intravenous drug use, HIV, or traumatic brain injury, were associated with atrophy or white matter diffusivity in the precariously housed sample.

## METHODS

### Study samples and participants

The Hotel Study is a prospective longitudinal observational study of individuals who are precariously housed in an impoverished neighbourhood of Vancouver, Canada.^10,13,14^ Participants were recruited from single-room occupancy hotels, a downtown community court, and a local hospital, and are assessed monthly and yearly on various aspects of health and functioning, including one or more multimodal MRI scans. Inclusion criteria included being aged 18 or older, ability to speak English, and ability to provide informed consent; no other exclusion criteria were applied. Participants in this study are demographically similar to studies on homelessness and have comparable health characteristics.^15,16^ Mental illness and substance use diagnoses were assessed using the Best Estimate Clinical Evaluation and Diagnosis Form by study psychiatrists,^17^ HIV status by baseline serology, and traumatic brain injury was operationalized as being struck in the head, neck, or face, and losing consciousness for any period of time, as assessed through a structured interview. Clinically significant findings were reported to the participants and their care providers. In the present study, we included baseline data from participants between the ages of 18 and 65 who had a usable T1-weighted and DTI scan (*n* = 312).

Data from the general population was collected as part of the population-based and open-access Cambridge Centre for Ageing and Neuroscience repository (CamCAN; http://www.mrc-cbu.cam.ac.uk/datasets/camcan/).^18,19^ The CamCAN dataset was selected as it is a comparably large dataset with all imaging acquired on a single MRI scanner. To facilitate comparison to the Hotel Study sample, we included participants who were between the age of 18 and 65 who had a usable T1-weighted and DTI scan (*n* = 382).

### Neuroimaging acquisition and processing

For the Hotel Study sample, scans were acquired on a 3T Philips Achieva scanner with an eight-channel SENSE head coil and for the CamCAN sample all scans were acquired on a 3T Siemens TIM Trio scanner with a 32-channel head coil. Scan acquisition parameters are detailed in the eMethods in the Supplement.

All scans were processed using similar processing pipelines. T1-weighted scans were processed using FreeSurfer version 6.0^20^ with full processing and quality control details outlined in the eMethods. Tissue-to-intracranial volume ratio was derived by dividing total cerebral brain tissue volume (excluding the ventricles, cerebellum, and brainstem) by estimated intracranial volume. Cortical thickness was extracted according to the Desikan-Killiany atlas^21^ and subcortical volumes were extracted according to the standard FreeSurfer subcortical segmentation atlas.^22^ Hotel Study DTI scans were processed using FSL version 5.0.11 and CamCAN DTI scans were processed with FSL version 6.0 (https://fsl.fmrib.ox.ac.uk),^23^ with full processing and quality control details outlined in the eMethods. Mean whole-brain fractional anisotropy (FA) and mean diffusivity (MD) data were extracted by taking an average from the white matter skeleton, and values across white matter tract regions of interest were exported according to the Johns Hopkins University white matter tract atlas.^24^

### Statistical analysis

We tested for differences between samples in median age with a Wilcoxon rank-sum test, and for differences in the proportion of sex between samples with a chi-square test. We centered all MRI measures in each sample to the sample mean (i.e., CamCAN values were centered on the CamCAN sample mean for that metric) and standardized the data. We report standardized beta weights throughout to facilitate comparison across imaging metrics and samples. We first looked at broad, whole-brain measures including tissue-to-intracranial volume ratio, cerebral white matter volume, cortical volume, subcortical grey matter volume, average whole-brain FA, and average whole-brain MD. We describe the difference in average slope between age and these whole-brain measures using multiple linear regression with a sample × age interaction term. However, there were non-linear associations between age and several whole-brain measures. Quadratic terms improperly describe non-linear relationships in cross-sectional studies^25^ and other non-linear methods (e.g., general additive models) yield coefficients that are challenging to interpret and compare across measures or samples. Therefore, we used piecewise regression to algorithmically estimate the age at which the breakpoints occurred and estimate slopes before and after the breakpoint.^26^ We also evaluated differential associations between age and cortical thickness, subcortical volumes, and white matter diffusivity across regions of interest using multiple linear regression with sample × age interaction terms. As shown in eFigure 1, linear approaches provided good estimates of average slope across the lifespan compared to non-linear approaches, so we used linear models to characterize region of interest results with one single standardized value for each region of interest. We covaried for estimated intracranial volume for all volumetric measures except for tissue-to-intracranial volume ratio and we covaried for sex for all measures.

**Figure 1.**
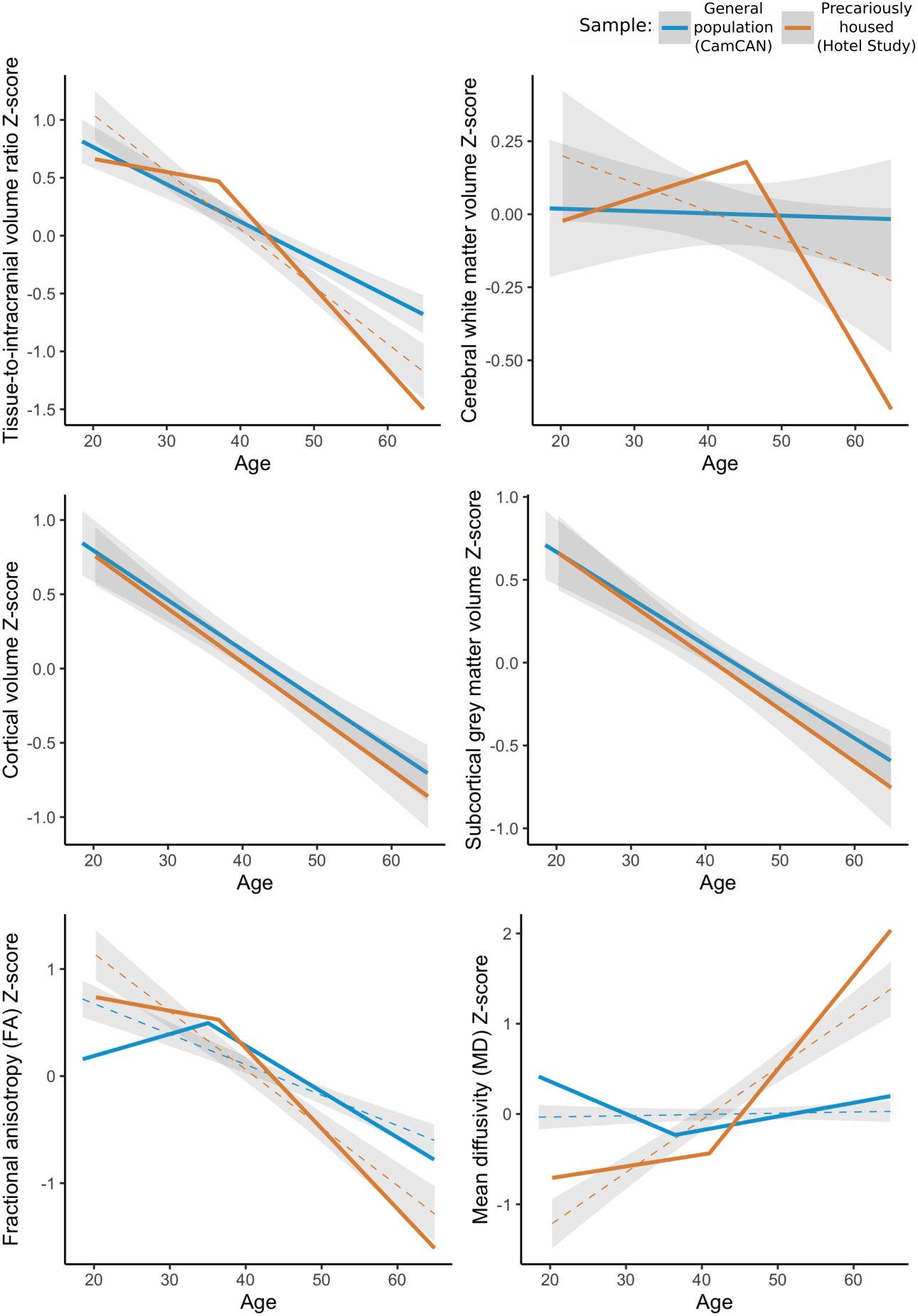
Whole-brain measures in the general population sample (CamCAN; blue) and in the precariously housed sample (Hotel Study; orange). Each metric is centered to the mean of the respective sample and standardized. For linear relationships, a linear fit and shaded 95% CI is shown. For non-linear relationships, the piecewise regression estimates are shown with a dashed linear (+ 95% CI) fit shown for reference.

One major factor that could disproportionately influence our results are individuals who had structural lesions due to traumatic brain injury, a subgroup which is overrepresented in the precariously housed sample compared to the general population.^27^ To evaluate whether our results were largely driven by these individuals, we first created a lesion overlap figure to describe the distribution and overlap of the lesions. We manually drew the lesion mask for each participant in FSL using T1-weighted and FLAIR images that were co-registered and then non-linearly registered the T1-weighted images and lesion masks to the MNI-152 standard space. To run the sensitivity analysis, we removed these participants and ran identical models to our original analyses.

Finally, we explored whether factors such as mental illness diagnoses, substance dependence, intravenous drug use, HIV positivity, or traumatic brain injury were associated with tissue-to-intracranial volume ratio or mean whole-brain FA in the precariously housed sample. To account for missing data we used multiple imputation as described in the eMethods. We used multiple linear regression with an age^2^ term to account for the non-linear relationships of the imaging variables with age, and then successively added blocks of variables.

All statistical analyses were conducted in R version 4.0.4,^28^ with piecewise regression implemented using the *segmented* package,^29^ region of interest results displayed using the *ggseg* package,^30^ and multiple imputation performed using the *mice* package.^31^

## RESULTS

The total sample included 694 participants, with 382 from the general population (CamCAN) sample and 312 from the precariously housed (Hotel Study) sample. The general population sample was slightly older and had a higher proportion of females.

The crude relationships between whole-brain imaging outcomes and age for each sample are shown in Figure 1, with adjusted estimates reported in-text and in eTable 1. Older age was associated with whole-brain atrophy in both samples; however, the precariously housed sample had a stronger association than the general population (*β*=-0.20, p=0.0029). The relationship between atrophy and age was linear in the general population and non-linear in the precariously housed sample, where older age was associated with more atrophy after age 37.0 (95% CI = 30.0–44.0). The association between age and atrophy was 6.5 times the slope before age 37 and approximately twice the magnitude of association as in the general population.

**Table 1.** Demographics of the study samples.

|  | **General population**<br>**(CamCAN; N =**<br>**382)** | **Precariously**<br>**housed (Hotel**<br>**Study; N = 312)** | **Test statistic** |
| --- | --- | --- | --- |
| **Age, median (IQR)** | 44.2 (33.5 - 54.8) | 41.8 (32.7 - 51.0) | $W = 67,022, p = 0.0047$ |
| **Sex, n/N males (%)** | 182/382 (47.6%) | 246/312 (78.8%) | $\chi^2 = 69.42, p < 0.0001$ |
| **Schizophrenia, n/N (%)** | — | 32/312 (10.3%) | — |
| **Schizoaffective, n/N (%)** | — | 33/312 (10.6%) | — |
| **Bipolar I or Bipolar NOS, n/N (%)** | — | 25/312 (8.0%) | — |
| **Bipolar II, n/N (%)** | — | 14/312 (4.5%) | — |
| **Major depressive disorder or depression NOS, n/N (%)** | — | 43/312 (13.8%) | — |
| **Psychotic disorder NOS, n/N (%)** | — | 33/312 (10.6%) | — |
| **Alcohol dependence, n/N (%)** | — | 60/312 (19.2%) | — |
| **Stimulant dependence, n/N (%)** | — | 243/312 (77.9%) | — |
| **Heroin dependence, n/N (%)** | — | 116/311 (37.3%) | — |
| **Cannabis dependence, *n/N* (%)** | — | 118/312 (37.8%) | — |
| **Intravenous drug use, *n/N* (%)** |  | 237/312 (76.0%) |  |
| **HIV, *n/N* (%)** | — | 39/288 (13.5%) | — |
| **Traumatic brain injury, *n/N* (%)** | — | 99/306 (32.4%) | — |

The differential atrophy in the precariously housed sample appeared to be largely driven by cerebral white matter volume (*β*=-0.12, *p*=0.0015), as neither cortical volume (*β*=-0.049, *p*=0.16) nor subcortical grey matter volume (*β*=-0.060, *p*=0.19) were significantly different in slope compared to the general population, and both declined linearly in each sample. The relationship between age and cerebral white matter volume was linear in the general population and stable across the age range. In contrast, the relationship was non-linear in the precariously housed sample as older age was associated with higher cerebral white matter volume up to age 39.8, after which it was associated with lower cerebral white matter volume.

There was also a stronger association between age and both whole-brain FA (*β*=-0.32, *p*<0.0001) and whole-brain MD (*β*=0.69, *p*<0.0001) in the precariously housed sample relative to the general population, with non-linear relationships in both samples for each metric. After age 36.4 in the precariously housed sample, older age was associated with lower FA at a magnitude 5.7 times that of the association before the breakpoint and approximately twice that of the general population at a similar age. After age 43.4 in the precariously housed sample, older age was associated with higher MD at a magnitude 5.4 times that of the association before the breakpoint and 7.9 times that of the general population at a similar age.

Differential associations with age for cortical thickness, subcortical volumes, and white matter tract diffusivity between the two samples is shown in Figure 2, with full results reported in eTables 2-4. In the precariously housed sample compared to the general population, older age was associated with lower cortical thickness in the middle and inferior temporal areas bilaterally, temporal poles, and the middle frontal and medial orbitofrontal areas, lower corpus callosum volume, higher third and lateral ventricle volumes, lower FA in 16 of 20 tracts, and higher MD in 14 of 20 tracts.

**Figure 2.**
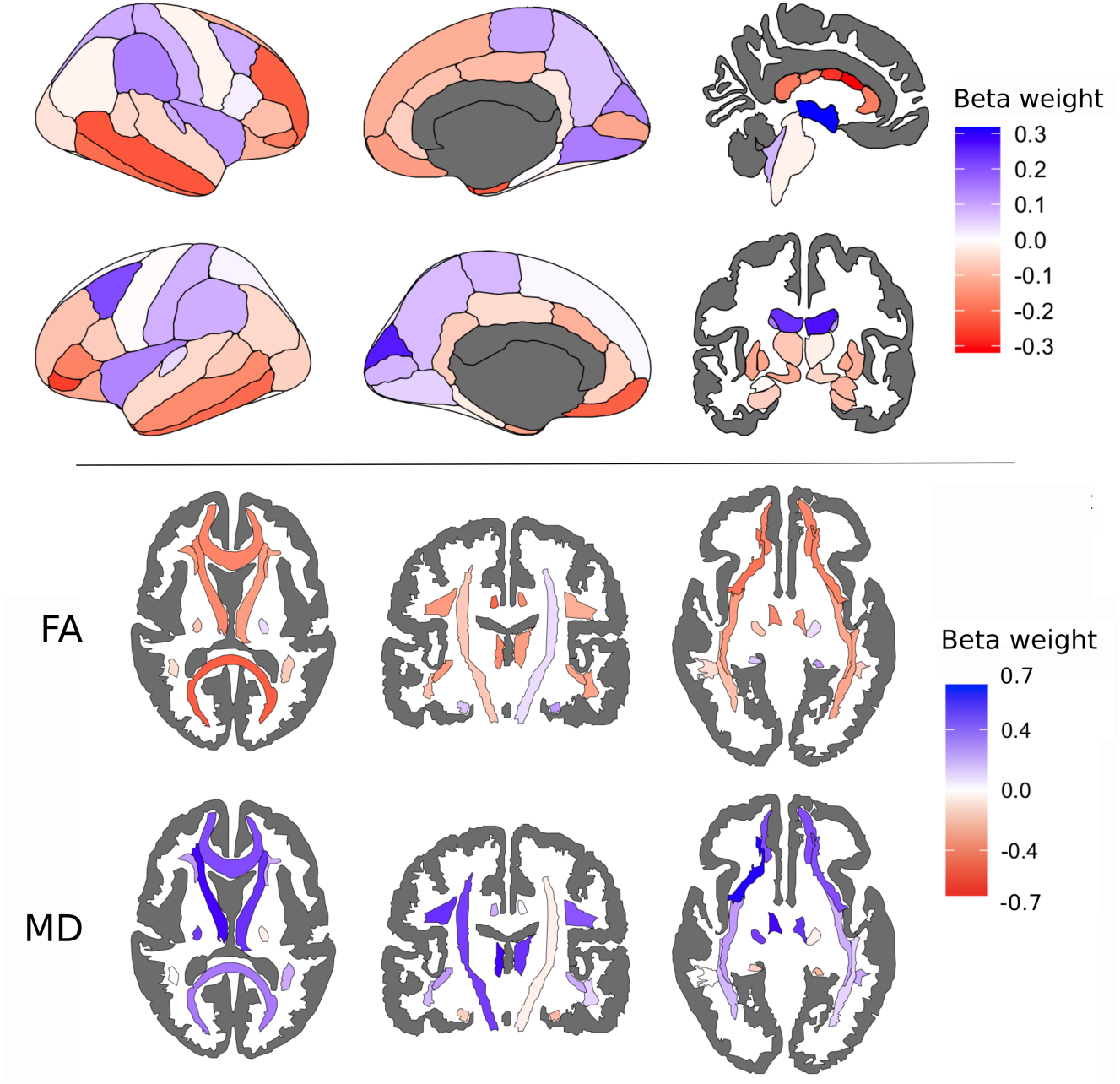
Differential associations across cortical, subcortical, and white matter regions of interest between the general population and precariously housed sample. Beta weight is the standardized beta from a sample × age interaction term for each region in the Desikan-Killiany atlas (top panel, left), FreeSurfer subcortical atlas (top panel, right), and John Hopkins University white matter atlas (bottom panel). Positive (blue) beta weights indicate that the relationship between the imaging metric and age is more positive in the precariously housed sample compared to the general population and negative (red) beta weights indicate that the relationship between the imaging metric and age is more negative in the precariously housed sample compared to the general population

Nineteen of the 312 precariously housed participants (6.1%) had MRI evidence of encephalomalacia attributable to traumatic brain injury, Figure 3. Traumatic brain injury lesions most commonly affected the frontal and orbitofrontal regions, as well as the temporal poles, consistent with the known vulnerability of these regions to trauma.^32^ There were no substantive changes in the nature or direction of our findings with these participants removed (eTable 5 and eFigures 2-4), indicating that our results are not unduly driven by these individuals.

**Figure 3.**
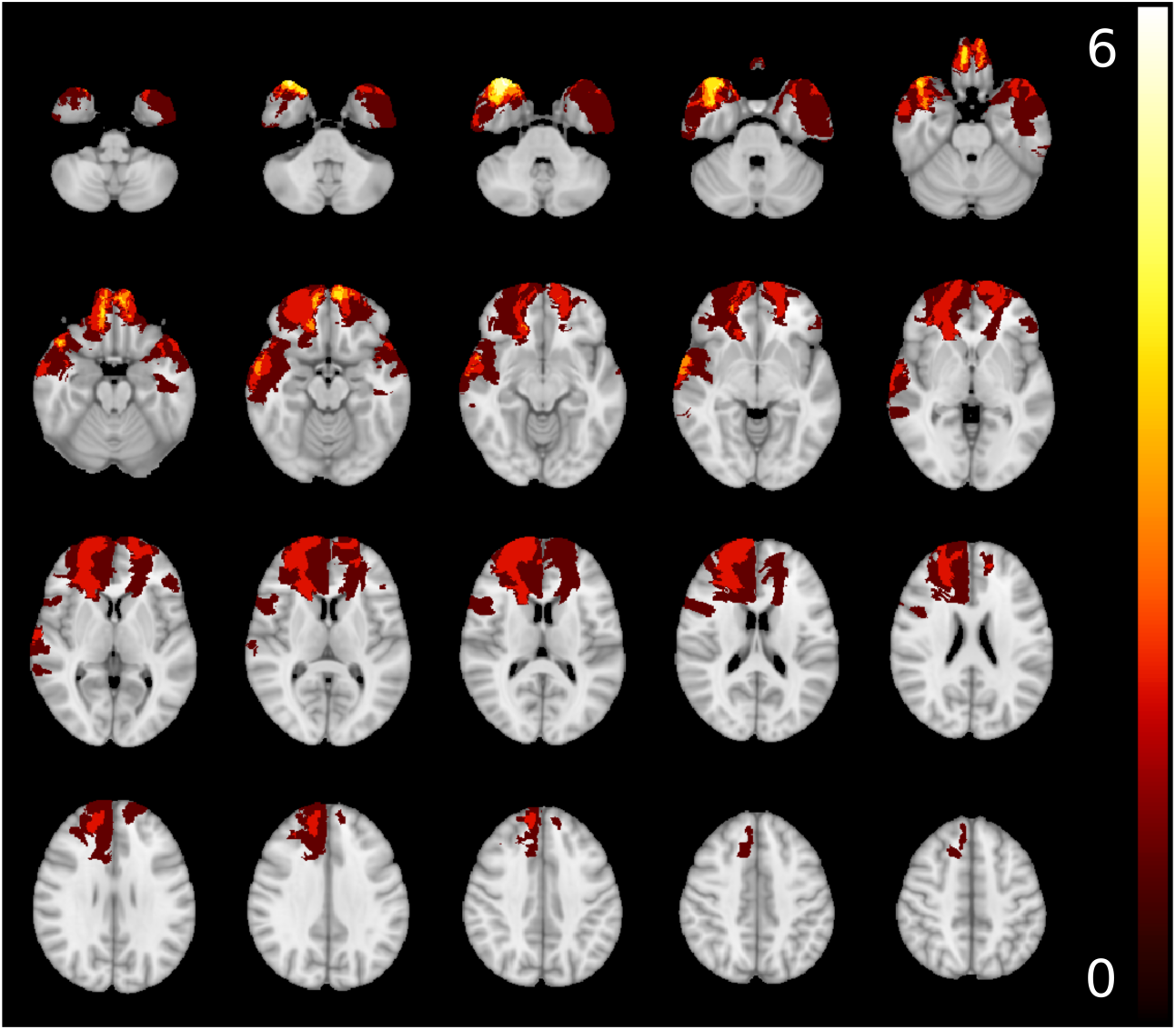
Overlap of lesions determined to be caused by traumatic brain injury in the precariously housed sample. Lesions are projected on the MNI-152 1mm template. Yellow/white denotes higher number of lesions overlapping.

Finally, we explored whether factors such as mental illness diagnoses, substance use, intravenous drug use, HIV, or history of traumatic brain injury were associated with atrophy or whole-brain FA in the precariously housed sample. The final blocks showing all risk factors we explored are shown in Figure 4 and the preliminary models and full results are reported in eTables 6-7. Male sex and history of traumatic brain injury were independently associated with greater atrophy. Heroin dependence was associated with lower FA and stimulant dependence was associated with higher FA.

**Figure 4.**
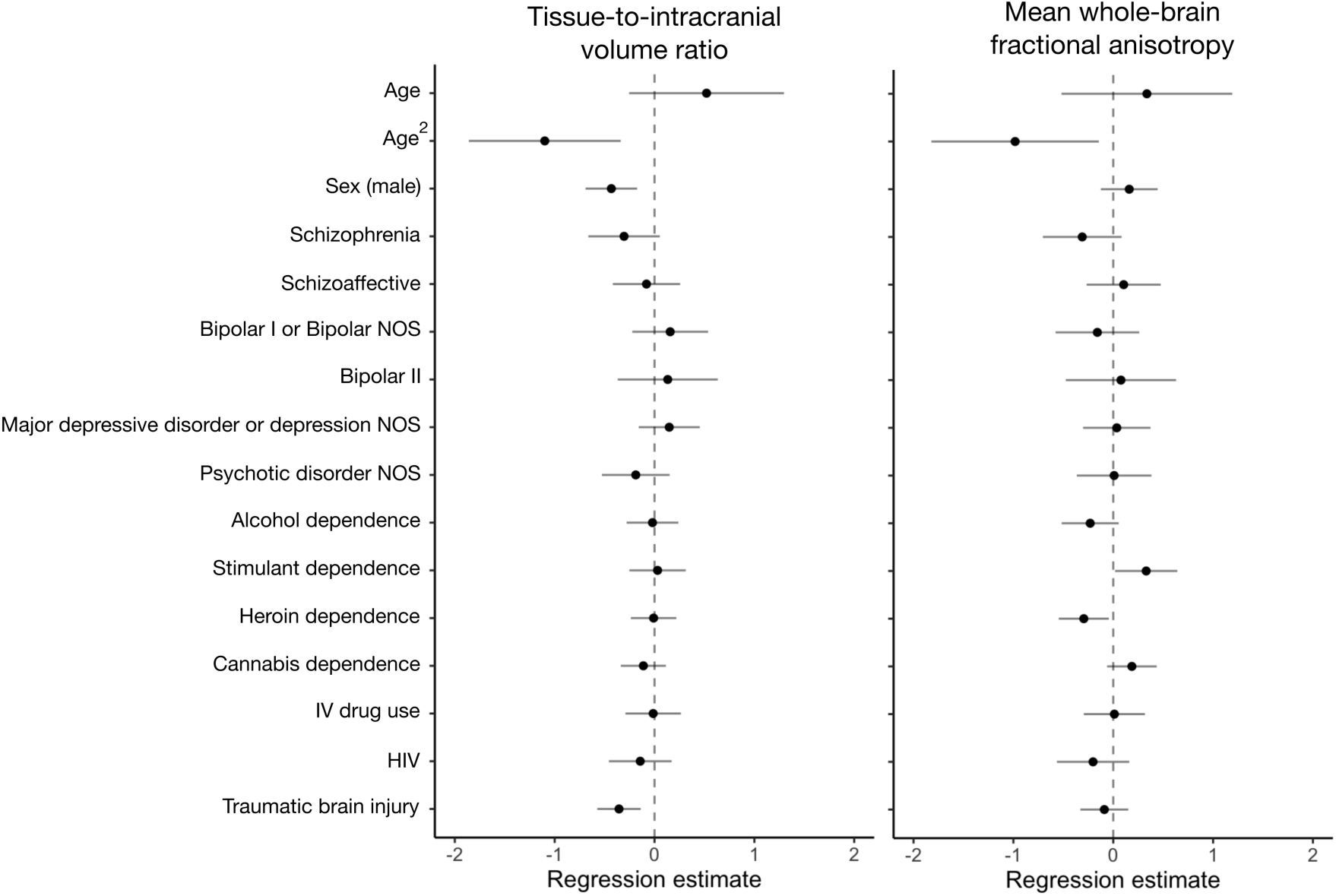
Forest plot of the associations between risk factors and tissue-to-intracranial volume (left) and risk factors and mean whole-brain fractional anisotropy (right). Error bars denote the 95% confidence interval for the regression estimate, and “NOS” denotes “not otherwise specified”.

## DISCUSSION

In this study we found that older age was associated with decreased MRI measures of brain structure among individuals who were homeless or precariously housed compared to the general population. We also found that male sex and history of traumatic brain injury were associated with more atrophy in the precariously housed sample, and that stimulant and heroin dependence were associated with altered white matter diffusivity.

The process of aging is generally understood to be complex and involves progressive allostatic load leading to physiologic and functional decline.^33,34^ The decline of MRI measures of brain structure is part of the normal course of aging in the general population and there is an accelerated decline in senescence.^35–38^ However, we show that homeless or precariously housed participants have an age-associated decline that occurs earlier and in greater magnitude of that seen in the general population. Our results most likely reflect the effects of both systemic factors (e.g., lack of housing) and individual factors (e.g., development of mental illness, bodily injury) that accumulate and interact across the lifespan. This accumulation may result in accelerated brain changes where MRI measures of brain structure decline earlier among homeless or precariously housed individuals, similar to the accelerated decline with senescence seen in older adults in the general population. This is supported by studies that have found a higher prevalence of other age-related conditions such as geriatric syndromes,^7,8^ multimorbidity,^10^ and earlier mortality.^2^

Our results could also be partly due to the development of pathological conditions that are distinct from normal aging. Brain atrophy, as assessed by structural neuroimaging, is a nonspecific biomarker for conditions such as Alzheimer’s disease^39,40^ and cerebral small vessel disease.^41,42^ Greater atrophy with age in our study may be indicative of a neurodegenerative process, especially among older participants. This is supported by previous research that has found that pathological conditions more often seen in older adults are also more common in this sample, with 46% having any neurological illness, 19% having a movement disorder, and 11% with a history of stroke.^9,10^

History of traumatic brain injury was associated with additional atrophy in the precariously housed sample which is congruent with studies that have found a range of neuroimaging changes, including atrophy, in individuals from the general population after traumatic brain injury.^43,44^ We also found that stimulant and heroin dependence were associated with altered whole-brain white matter diffusivity. Previous studies report mixed findings and that the direction of the association between substance use and structural MRI measures depends on the specific substance and the region of interest.^45,46^

Our work has potential implications for care providers, policy makers, and researchers. First, care providers should be aware of this differential decline in MRI measures of brain structure and consider earlier evaluation and intervention for individuals experiencing homelessness or precarious housing. History of traumatic brain injury and substance dependence represent especially important factors to consider, and our work highlights the importance of accurate medical and injury history when working with vulnerable groups, especially given the association between traumatic brain injury and homelessness.^6,47^ Second, our study contributes to the growing evidence showing a high burden of functional impairment, morbidity, and mortality among these individuals. Policymakers should consider increasing targeted service availability, or expanding service eligibility; for example, making supportive services targeted for seniors in the general population available to individuals in these groups at a younger age. Finally, future research should characterize predictors of longitudinal change in these groups and evaluate how changes in brain structure is associated with functioning. Age-related decline in cognitive function, for example, is associated with adverse changes in MRI measures of brain structure,^36^ and thus, our results likely partially explain the well-documented cognitive and functional impairments experienced by these individuals.^48,49^ Intervention studies specifically informed by the unique health challenges of this population (e.g., earlier targeted cerebrovascular risk factor management strategies including non-traditional risk factors such as IV drug use^9^) are also warranted in an effort to improve overall health.

## Limitations

While our study leverages a well-characterized sample of precariously housed individuals and a large open-access sample from the general population, our work has several limitations. First, the two samples were acquired on different scanners with different acquisition parameters, and thus, comparison of the intercepts or “main effects” could be biased by differences between scanners or acquisitions.^50^ We restricted our analyses to look only at the difference in slope between samples, however, imaging metrics could be different, on average, in the precariously housed sample compared to the general population (i.e., without a difference in slope) which we were unable to cogently evaluate in this study. Second, while quantitative MRI has been used extensively to evaluate brain structure *in vivo*, it is important to acknowledge that MRI measures of brain structure may be biased by a variety of factors and are not fully representative of the underlying biology.^51^ Finally, we restricted our exploratory analyses on predictors of atrophy and FA to whole-brain measures. It is possible that some factors are associated with alterations in specific regions of interest but not with whole-brain measures. Future studies are needed to characterize the focal brain changes associated with these risk factors.

## Conclusions

Older age is associated with adverse MRI measures of brain structure among individuals who are homeless or precariously housed well beyond the normal pattern seen in the general population. History of traumatic brain injury and substance dependence are factors that are overrepresented among these individuals and associated with further atrophy and alterations in white matter diffusivity. Changes in care provision and policy are needed to address the disparity in health outcomes for individuals experiencing homelessness and precarious housing.

## Data Availability

Data from the Cambridge Centre for Aging and Neuroscience is available at: https://www.cam-can.org. Data from the Hotel Study cannot be made publicly available due to possible privacy breaches and other ethical and legal obligations to the study participants. These restrictions are outlined by the University of British Columbia Clinical Research Ethics Board and the Simon Fraser University Research Ethics Board. Inquiries regarding data can be made to the Clinical Research Ethics Board of the University of British Columbia and the study principal investigator (WGH).

## ACKNOWLEDGEMENTS

Funding support for the Hotel Study includes the Canadian Institutes of Health Research (MOP-137103, MOP-390996, PJT-169094) and the BC Mental Health and Substance Use Services Research Institute. Data collection and sharing for this project for the CamCAN sample was provided by the Cambridge Centre for Ageing and Neuroscience. CamCAN funding was provided by the UK Biotechnology and Biological Sciences Research Council (grant number BB/H008217/1), together with support from the UK Medical Research Council and the University of Cambridge, UK.

