## Supplementary materials for "Differential age-associated brain atrophy and white matter changes among homeless and precariously housed individuals compared to the general population"

### Supplementary Material

**eMethods**
**eFigure 1**. Predicted values of tissue-to-intracranial volume ratio as a function of age in the Hotel Study as estimated by three different methods.
**eTable 1**. Full adjusted results of whole-brain imaging outcomes.
**eTable 2**. Full results of cortical thickness across regions of interest.
**eTable 3**. Full results of subcortical volume across regions of interest.
**eTable 4**. Full results of white matter diffusivity across regions of interest.
**eTable 5**. Sensitivity analysis on whole-brain measures.
**eFigure 2**. Cortical regions of interest results with participants who had MRI evidence of traumatic brain injury removed.
**eFigure 3**. Subcortical regions of interest results with participants who had MRI evidence of traumatic brain injury removed.
**eFigure 4**. White matter tract regions of interest with participants who had MRI evidence of traumatic brain injury removed.
**eTable 6**. Full results of factors associated with tissue-to-intracranial volume ratio in the precariously housed sample.
**eTable 7**. Full results of factors associated with whole-brain fractional anisotropy (FA) in the precariously housed sample
**eReferences**

### eMethods

For the Hotel Study sample, all scans were acquired using 3T Philips Achieva (software version 2.6.3.5) using an eight-channel SENSE head coil. High resolution 3D T1-weighted FFE sagittal images were acquired with TE = 3.7 ms, TR = 8.1 ms, flip angle 8°, FOV = 256mm × 256mm, acquisition matrix = 256 × 250, reconstruction matrix = 256 × 256, voxel spacing = 1.0 mm × 1.0 mm × 1.0 mm, 190 contiguous slices, slice thickness = 1 mm, gap = 0, SENSE = 1, scan duration = 7:23 min. Diffusion-weighted images were acquired during the same session with 70 contiguous transverse slices in 32 directions, acquisition matrix = 100 × 99, reconstruction matrix = 112 × 112, acquisition voxel = 2.24 × 2.24 × 2.0 mm, reconstructed voxel = 2.0 × 2.0 × 2.0 mm, slice thickness = 2.2 mm, TE = 60 ms, TR = 6451 ms, FOV = 224 mm × 224 mm, flip angle = 90°, SENSE = 2.1, maximum number of gradient orientations = 33, $b$ = 700 s/mm$^2$, total scan duration = 3:46 min.

For the CamCAN sample, all scans were acquired on a 3T Siemens TIM Trio scanner with a 32-channel head coil. T1-weighted scans were acquired using an MPRAGE sequence, with TR = 2250ms, TE = 2.99ms, flip angle = 9°, FOV = 256 × 240 × 192mm, voxel size = 1 × 1 × 1mm, and GRAPPA 2.[1] Diffusion-weighted scans were acquired with a twice-refocused SE sequence at three $b$-values ($b$ = 1000, 30 directions, 66 axial slices; $b$ = 2000, 30 directions, 66 axial slices, and $b$ = 0, 66 axial slices, 3 images), with TR = 9100ms, TE = 104ms, FOV = 192 × 192mm, voxel size = 2 × 2 × 2mm.

T1-weighted data from both samples was processed using FreeSurfer version 6.0 (https://surfer.nmr.mgh.harvard.edu)[2] and involved removal of non-brain tissue using a hybrid watershed/surface deformation procedure,[3] automated Talairach transformation and segmentation of the subcortical white matter and deep gray matter structures,[4,5] intensity normalization,[6] tessellation of the gray-white matter boundary, automated topology correction,[7,8] and surface deformation following intensity gradients to optimally place gray-white and gray-cerebrospinal fluid boundaries.[9–11] T1-weighted data for Hotel Study participants was evaluated by a trained research assistant and edited where necessary due to the higher pathology than the general population. T1-weighted data in the CamCAN sample was quality controlled using the ENIGMA Consortium quality control protocol (https://enigma.usc.edu/).

Diffusion tensor imaging scans for both study samples were processed using FSL version 6.0 (https://fsl.fmrib.ox.ac.uk),[12] which involved brain extraction,[13] eddy current correction,[14] and tensor fitting. Additionally, for the Hotel Study scans, *eddy_cuda8.0* was used to detect and correct slices that were corrupted by motion-induced signal dropout including slice-to-volume registration. Non-diffusion volume and intensity inversed T1 volumes were used to estimate susceptibility distortion by using ANT's SyN algorithm,[15] and then mapped to T1 space and resliced as 2mm × 2mm × 2mm isotropically. All DTI volumes were then processed in FSL with tract-based spatial statistics,[16] which involved aligning individual-participant FA data into a common space using nonlinear registration, creating a mean FA image, thinning the mean FA image it into a white matter skeleton, and projecting each participants' aligned data onto the white matter skeleton.[16] Quality control was performed by a trained research assistant on the raw data, after eddy current correction, and after tensor fitting.

There was a relatively small amount of missing data for our analyses of factors associated with tissue-to-intracranial volume ratio and mean whole-brain fractional anisotropy in the precariously housed sample. HIV status was missing for 7.69%, traumatic brain injury history for 1.92%, IV drug use for 0.64%, and heroin dependence for 0.32%. To retain all participants in the analyses we used multiple imputation implemented through the *mice* package in R.[17,18] We used all variables included in the analyses (age, sex, tissue-to-intracranial volume ratio, mean whole-brain fractional anisotropy, mental illness diagnoses, substance dependence diagnoses, IV drug use, HIV status, and traumatic brain injury history) to impute 10 new datasets. Each regression model was fit to each imputed dataset and the results were pooled to yield the final estimates. There were no differences in the direction or statistical significance of the results between the results based on the imputed data and identical models fit on the observed data, therefore the imputed results are reported in-text and in the supplement.

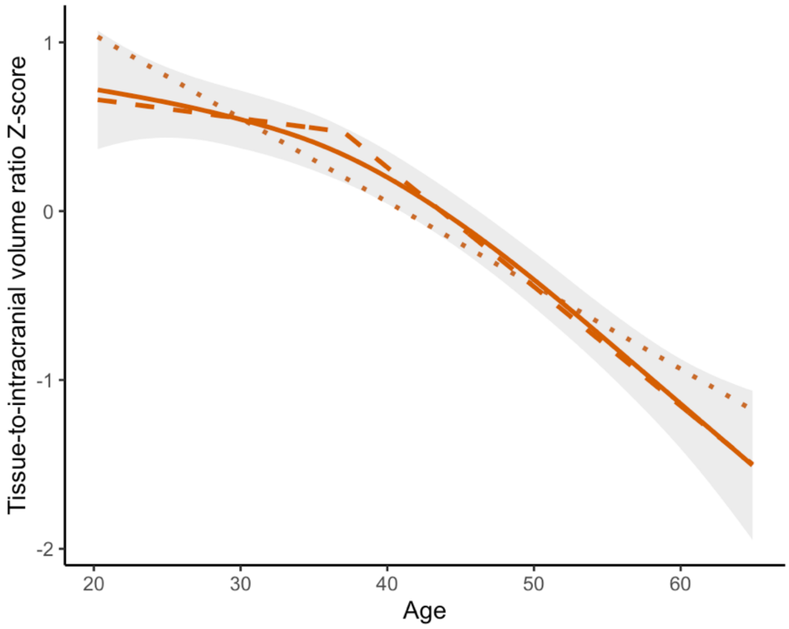

**eFigure 1. Predicted values of tissue-to-intracranial volume ratio as a function of age in the Hotel Study as estimated by three different methods.** Linear regression (dotted line; adjusted $R^2$ = 0.265), piecewise linear regression (dashed line; adjusted $R^2$ = 0.283), and general additive model (solid line with 95% confidence interval in shaded region; adjusted $R^2$ =0.282).

### eTable 1. Full adjusted results of whole-brain imaging outcomes.

| Metric | Sample | Beta[1] | Within-group slope estimates (with break points if applicable) |
|---|---|---|---|
| Tissue-to-intracranial volume ratio[2] | General population (CamCAN) | β = -0.20, p = 0.0029 | Linear: -0.39 |
| | Precariously housed (Hotel Study) | | Piecewise: -0.13 up to age 37, then -0.88 |
| Cerebral white matter volume[3] | General population (CamCAN) | β = -0.12, p = 0.0015 | Linear: 0.038 |
| | Precariously housed (Hotel Study) | | Piecewise: 0.23 up to age 39.8, then -0.33 |
| Cortical grey matter volume[3] | General population (CamCAN) | β = -0.049, p = 0.16 | Linear: -0.41 |
| | Precariously housed (Hotel Study) | | Linear: -0.36 |
| Subcortical grey matter volume[3] | General population (CamCAN) | β = -0.060, p = 0.19 | Linear: -0.35 |
| | Precariously housed (Hotel Study) | | Linear: -0.30 |
| Fractional anisotropy (FA)[2] | General population (CamCAN) | β = -0.32, p < 0.0001 | Piecewise: 0.59 up to age 30.6, then -0.47 |
| | Precariously housed (Hotel Study) | | Piecewise: -0.16 up to age 36.4, then -0.91 |

| | | | |
|---|---|---|---|
| Mean diffusivity (MD)[2] | General population (CamCAN) | β = 0.69 p < 0.0001 | Piecewise: -0.44 up to age 35.9, then 0.17 |
| | Precariously housed (Hotel Study) | | Piecewise: 0.25 up to age 43.4, then 1.34 |

[1]Standardized beta of the sample x age interaction term; [2]adjusted for sex; [3]adjusted for sex and intracranial volume

### eTable 2. Full results of cortical thickness across regions of interest.

| Region of interest | Beta[1] | p-value |
|---|---|---|
| Banks of the superior temporal sulcus (left) | -6.50E-02 | 0.38 |
| Banks of the superior temporal sulcus (right) | -5.03E-02 | 0.49 |
| Caudal anterior cingulate (left) | -9.94E-02 | 0.18 |
| Caudal anterior cingulate (right) | -7.83E-02 | 0.29 |
| Caudal middle frontal (left) | -1.94E-01 | 0.0072 |
| Caudal middle frontal (right) | 8.39E-02 | 0.25 |
| Cuneus (left) | 2.48E-01 | 0.0011 |
| Cuneus (right) | 1.25E-01 | 0.1 |
| Entorhinal (left) | -1.14E-01 | 0.14 |
| Entorhinal (right) | -2.10E-01 | 0.007 |
| Frontal pole (left) | -5.76E-02 | 0.46 |
| Frontal pole (right) | -1.51E-02 | 0.84 |
| Fusiform (left) | -6.54E-03 | 0.93 |
| Fusiform (right) | -1.75E-02 | 0.82 |
| Inferior parietal (left) | -4.83E-02 | 0.5 |
| Inferior parietal (right) | -1.32E-02 | 0.85 |
| Inferior temporal (left) | -1.90E-01 | 0.011 |
| Inferior temporal (right) | -1.89E-01 | 0.012 |
| Insula (left) | 1.27E-01 | 0.086 |
| Insula (right) | 1.07E-01 | 0.14 |
| Isthmus cingulage (left) | -6.42E-01 | 0.39 |
| Isthmus cingulage (right) | -2.92E-02 | 0.69 |
| Lateral occipital (left) | -5.64E-02 | 0.46 |
| Lateral occipital (right) | -3.71E-02 | 0.63 |
| Lateral orbitofrontal (left) | -1.09E-01 | 0.13 |
| Lateral orbitofrontal (right) | -9.97E-02 | 0.18 |
| Lingual (left) | 4.64E-02 | 0.53 |
| Lingual (right) | 1.37E-01 | 0.067 |
| Medial orbitofrontal (left) | -2.04E-01 | 0.0067 |
| Medial orbitofrontal (right) | -1.26E-01 | 0.093 |
| Middle temporal (left) | -1.49E-01 | 0.033 |
| Middle temporal (right) | -2.16E-01 | 0.002 |
| Paracentral (left) | 7.12E-02 | 0.34 |
| Paracentral (right) | 8.99E-02 | 0.22 |
| Parahippocampal (left) | -2.33E-02 | 0.76 |
| Parahippocampal (right) | 4.57E-03 | 0.95 |
| Pars opercularis (left) | -6.40E-02 | 0.35 |
| Pars opercularis (right) | 1.60E-02 | 0.82 |
| Pars orbitalis (left) | -2.45E-01 | 0.001 |
| Pars orbitalis (right) | -1.66E-01 | 0.025 |
| Pars triangularis (left) | -1.61E-01 | 0.019 |
| Pars triangularis (right) | -8.26E-02 | 0.23 |
| Pericalcarine (left) | 7.41E-02 | 0.33 |
| Pericalcarine (right) | -1.15E-01 | 0.14 |
| Postcentral (left) | 7.92E-01 | 0.28 |
| Postcentral (right) | 7.71E-02 | 0.3 |
| Posterior cingulate (left) | -4.49E-02 | 0.53 |

| | | |
|---|---|---|
| Posterior cingulate (right) | -8.63E-02 | 0.23 |
| Precentral (left) | -4.91E-03 | 0.95 |
| Precentral (right) | -1.41E-02 | 0.84 |
| Precuneus (left) | 6.62E-02 | 0.35 |
| Precuneus (right) | 6.30E-02 | 0.38 |
| Rostral anterior cingulate (left) | -6.01E-02 | 0.4 |
| Rostral anterior cingulate (right) | -6.07E-02 | 0.42 |
| Rostral middle frontal (left) | -7.64E-02 | 0.3 |
| Rostral middle frontal (right) | -2.04E-01 | 0.0052 |
| Superior frontal (left) | 8.16E-03 | 0.9 |
| Superior frontal (right) | -1.02E-01 | 0.14 |
| Superior parietal (left) | 1.10E-02 | 0.88 |
| Superior parietal (right) | 8.43E-02 | 0.25 |
| Superior temporal (left) | -4.93E-02 | 0.48 |
| Superior temporal (right) | -5.43E-02 | 0.44 |
| Supramarginal (left) | 8.03E-02 | 0.25 |
| Supramarginal (right) | 1.30E-01 | 0.06 |
| Temporal pole (left) | -2.34E-01 | 0.0027 |
| Temporal pole (right) | -2.49E-01 | 0.0014 |
| Transverse temporal (left) | 4.02E-02 | 0.59 |
| Transverse temporal (right) | 8.51E-02 | 0.26 |

[1]Standardized beta of the sample × age interaction term

### eTable 3. Full results of subcortical volume across regions of interest.

| Region of interest | Beta[1] | p-value |
|---|---|---|
| 3rd ventricle | 3.17E-01 | < 0.0001 |
| 4th ventricle | 8.21E-02 | 0.26 |
| Amygdala (left) | -8.17E-03 | 0.89 |
| Amygdala (right) | -9.76E-02 | 0.1 |
| Brainstem | -1.66E-02 | 0.77 |
| Caudate (left) | 1.45E-01 | 0.024 |
| Caudate (right) | 1.04E-01 | 0.1 |
| Corpus callosum (anterior) | -1.75E-01 | 0.013 |
| Corpus callosum (central) | -2.71E-01 | 0.00018 |
| Corpus callosum (mid-anterior) | -3.19E-01 | < 0.0001 |
| Corpus callosum (mid-posterior) | -1.50E-01 | 0.041 |
| Corpus callosum (posterior) | -1.83E-01 | 0.012 |
| Hippocampus (left) | -6.43E-02 | 0.32 |
| Hippocampus (right) | -8.63E-02 | 0.18 |
| Lateral ventricle (left) | 2.40E-01 | 0.00028 |
| Lateral ventricle (right) | 2.70E-01 | < 0.0001 |
| Pallidum (left) | -1.21E-01 | 0.043 |
| Pallidum (right) | -1.15E-01 | 0.053 |
| Putamen (left) | -1.21E-01 | 0.046 |
| Putamen (right) | -1.04E-01 | 0.079 |
| Thalamus (left) | -7.33E-02 | 0.17 |
| Thalamus (right) | -2.25E-02 | 0.66 |
| VentralDC (left) | -1.15E-01 | 0.053 |
| VentralDC (right) | -5.75E-02 | 0.32 |

[1]Standardized beta of the sample × age interaction term

### eTable 4. Full results of white matter diffusivity across regions of interest.

| | Fractional anisotropy (FA) | | Mean diffusivity (MD) | | Axial diffusivity (AD) | | Radial diffusivity (RD) | |
|---|---|---|---|---|---|---|---|---|
| **Region of interest** | **Beta[1]** | **p-value** | **Beta[1]** | **p-value** | **Beta[1]** | **p-value** | **Beta[1]** | **p-value** |
| Anterior thalamic radiation (left) | -3.50E-01 | < 0.0001 | 6.10E-01 | < 0.0001 | 5.50E-01 | < 0.0001 | 6.20E-01 | < 0.0001 |
| Anterior thalamic radiation (right) | -2.90E-01 | < 0.0001 | 5.20E-01 | < 0.0001 | 4.50E-01 | < 0.0001 | 5.40E-01 | < 0.0001 |
| Cingulate gyrus (left) | -4.60E-01 | < 0.0001 | 1.90E-01 | 0.01 | -3.30E-01 | < 0.0001 | 4.70E-01 | < 0.0001 |
| Cingulate gyrus (right) | -3.40E-01 | < 0.0001 | 1.50E-02 | 0.85 | -3.50E-01 | < 0.0001 | 3.20E-01 | < 0.0001 |
| Hippocampus (left) | 1.40E-01 | 0.075 | -1.40E-01 | 0.075 | -9.60E-02 | 0.21 | -1.50E-01 | 0.045 |
| Hippocampus (right) | 2.30E-01 | 0.0025 | -2.10E-01 | 0.0067 | -8.70E-02 | 0.25 | -2.50E-01 | 0.0012 |
| Corticospinal tract (left) | -1.60E-01 | 0.028 | 4.90E-01 | < 0.0001 | 3.20E-01 | < 0.0001 | 4.10E-01 | < 0.0001 |
| Corticospinal tract (right) | 7.90E-02 | 0.3 | -4.30E-02 | 0.58 | -1.40E-02 | 0.85 | -5.20E-02 | 0.5 |
| Forceps major | -4.80E-01 | < 0.0001 | 3.30E-01 | < 0.0001 | -1.90E-01 | 0.014 | 4.90E-01 | < 0.0001 |
| Forceps minor | -3.70E-01 | < 0.0001 | 4.50E-01 | < 0.0001 | 6.00E-02 | 0.44 | 5.30E-01 | < 0.0001 |
| Inferior fronto-occipital fasciculus (left) | -3.00E-01 | < 0.0001 | 2.40E-01 | 0.0016 | -8.20E-02 | 0.29 | 3.80E-01 | < 0.0001 |
| Inferior fronto-occipital fasciculus (right) | -2.70E-01 | 0.00011 | 1.90E-01 | 0.01 | -1.60E-01 | 0.036 | 3.60E-01 | < 0.0001 |
| Inferior longitudinal fasciculus (left) | -1.90E-01 | 0.0088 | 1.60E-01 | 0.034 | 9.20E-03 | 0.91 | 2.60E-01 | 0.00059 |
| Inferior longitudinal fasciculus (right) | -2.70E-01 | 0.00014 | 1.00E-01 | 0.18 | -1.90E-01 | 0.012 | 2.80E-01 | 0.00015 |
| Superior longitudinal fasciculus (left) | -3.00E-01 | < 0.0001 | 5.30E-01 | < 0.0001 | 3.70E-01 | < 0.0001 | 5.00E-01 | < 0.0001 |
| Superior longitudinal fasciculus (right) | -2.40E-01 | 0.00072 | 4.20E-01 | < 0.0001 | 1.90E-01 | 0.012 | 4.50E-01 | < 0.0001 |
| Uncinate fasciculus (left) | -4.10E-01 | < 0.0001 | 6.70E-01 | < 0.0001 | 2.90E-01 | 0.00023 | 6.40E-01 | < 0.0001 |
| Uncinate fasciculus (right) | -3.70E-01 | < 0.0001 | 4.50E-01 | < 0.0001 | 7.80E-02 | 0.32 | 4.90E-01 | < 0.0001 |
| Superior longitudinal fasciculus temporal portion (left) | -9.60E-02 | 0.2 | 2.70E-03 | 0.97 | -1.10E-01 | 0.15 | 9.00E-02 | 0.24 |
| Superior longitudinal fasciculus | -1.50E-01 | 0.047 | 2.00E-01 | 0.0094 | 1.60E-02 | 0.84 | 2.50E-01 | 0.0013 |

| |
|---|
| temporal portion (right) |

[1]Standardized beta of the interaction term

**eTable 5. Sensitivity analysis on whole-brain measures.**

| Metric | Sample | Beta[1] with all participants included | Beta[1] with participants who had MRI evidence of traumatic brain injury removed |
|---|---|---|---|
| Tissue-to-intracranial volume ratio[1] | General population (CamCAN) | β = -0.20, p = 0.0029 | β = -0.18, p = 0.013 |
| | Precariously housed (Hotel Study) | | |
| Cerebral white matter volume[2] | General population (CamCAN) | β = -0.12, p = 0.0015 | β = -0.10, p = 0.010 |
| | Precariously housed (Hotel Study) | | |
| Cortical grey matter volume[2] | General population (CamCAN) | β = -0.049, p = 0.16 | β = -0.044, p = 0.22 |
| | Precariously housed (Hotel Study) | | |
| Subcortical grey matter volume[2] | General population (CamCAN) | β = -0.060, p = 0.19 | β = -0.045, p = 0.34 |
| | Precariously housed (Hotel Study) | | |
| Fractional anisotropy (FA)[1] | General population (CamCAN) | β = -0.32, p < 0.0001 | β = -0.32, p < 0.0001 |
| | Precariously housed (Hotel Study) | | |
| Mean diffusivity (MD)[1] | General population (CamCAN) | β = 0.69 p < 0.0001 | β = 0.68, p < 0.0001 |
| | Precariously housed (Hotel Study) | | |

[1]Standardized beta of the sample x age interaction term; [2]adjusted for sex; [3]adjusted for sex and intracranial volume

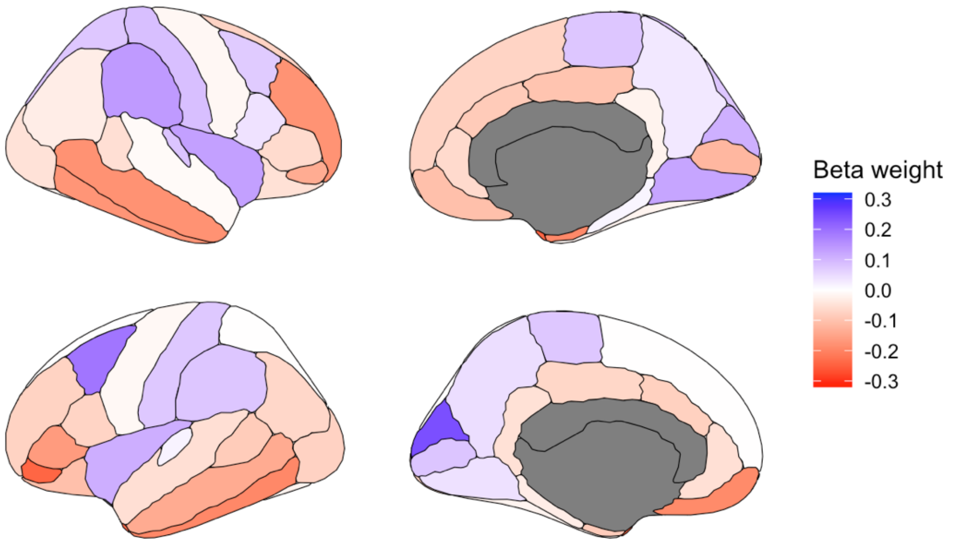

**eFigure 2. Cortical regions of interest results with participants who had MRI evidence of traumatic brain injury removed.**

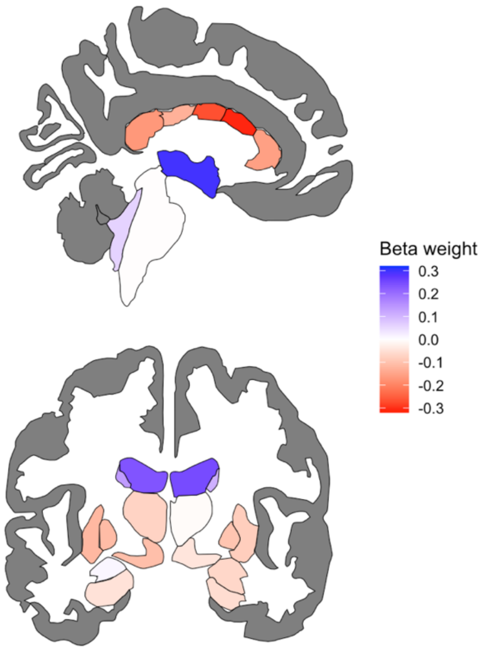

**eFigure 3. Subcortical regions of interest results with participants who had MRI evidence of traumatic brain injury removed.**

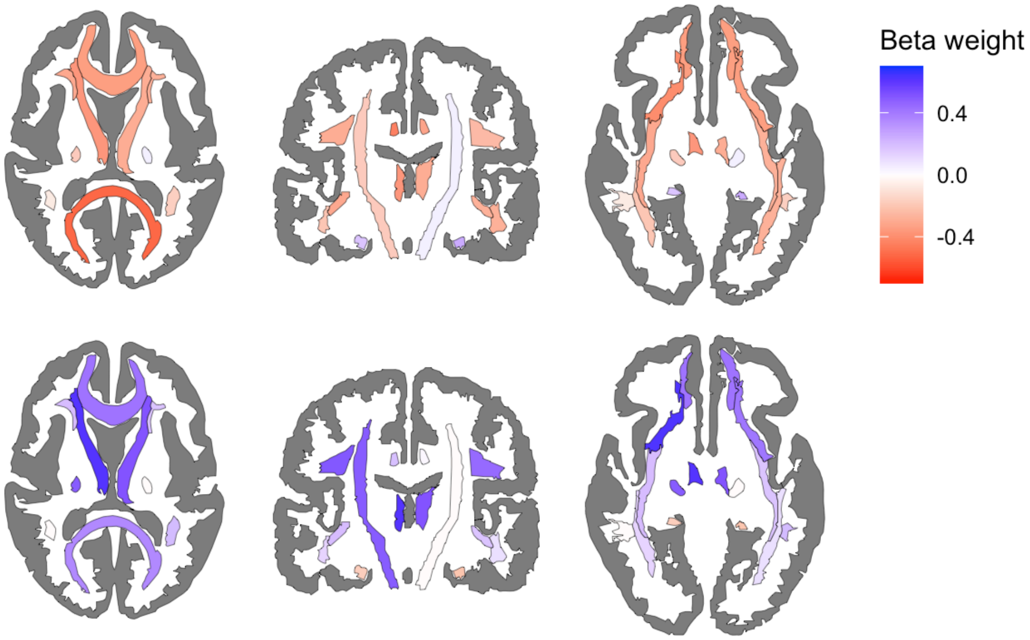

**eFigure 4. White matter tract regions of interest results with participants who had MRI evidence of traumatic brain injury removed.**

**eTable 6. Full results of factors associated with tissue-to-intracranial volume ratio in the precariously housed sample**.

| **Tissue-to-intracranial volume ratio** | **Block 1: Basic demographics** | | | **Block 2: Mental health diagnoses** | | | **Block 3: Substance use** | | | **Block 4: HIV and traumatic brain injury** | | |
|---|---|---|---|---|---|---|---|---|---|---|---|---|
| | **Beta** | **95% CI** | ***p*-value** | **Beta** | **95% CI** | ***p*-value** | **Beta** | **95% CI** | ***p*-value** | **Beta** | **95% CI** | ***p*-value** |
| Age | 0.56 | -0.15, 1.27 | 0.12 | 0.55 | -0.19, 1.28 | 0.14 | 0.49 | -0.30, 1.27 | 0.23 | 0.52 | -0.25, 1.30 | 0.19 |
| $Age^2$ | -1.12 | -1.83, -0.40 | 0.0022 | -1.13 | -1.86, -0.40 | 0.0027 | -1.08 | -1.85, -0.31 | 0.0062 | -1.09 | -1.86, -0.34 | 0.0048 |
| Sex | | | | | | | | | | | | |
| Female | — | — | | — | — | | — | — | | — | — | |
| Male | -0.51 | -0.75, -0.27 | < 0.0001 | -0.45 | -0.70, -0.17 | 0.00047 | -0.43 | -0.70, -0.17 | 0.0013 | -0.43 | -0.69, -0.17 | 0.0011 |
| Schizophrenia | | | | -0.29 | -0.65, 0.06 | 0.11 | -0.31 | -0.67, 0.05 | 0.096 | -0.31 | -0.66, 0.05 | 0.095 |
| Schizoaffective | | | | -0.05 | -0.39, 0.28 | 0.76 | -0.05 | -0.39, 0.29 | 0.78 | -0.08 | -0.42, 0.26 | 0.64 |
| Bipolar I or Bipolar NOS | | | | 0.11 | -0.28, 0.48 | 0.6 | 0.11 | -0.27, 0.50 | 0.56 | 0.16 | -0.22, 0.54 | 0.42 |
| Bipolar II | | | | 0.12 | -0.37, 0.62 | 0.63 | 0.13 | -0.38, 0.63 | 0.62 | 0.13 | -0.37, 0.63 | 0.6 |
| Major depressive disorder or depression NOS | | | | 0.16 | -0.14, 0.46 | 0.31 | 0.16 | -0.15, 0.47 | 0.32 | 0.15 | -0.16, 0.45 | 0.34 |
| Psychotic disorder NOS | | | | -0.21 | -0.55, 0.12 | 0.22 | -0.22 | -0.56, 0.13 | 0.21 | -0.19 | -0.53, 0.15 | 0.28 |
| Alcohol | | | | | | | -0.01 | -0.27, 0.25 | 0.94 | -0.02 | -0.28, 0.24 | 0.88 |
| Stimulant | | | | | | | 0.07 | -0.22, 0.35 | 0.65 | 0.03 | -0.25, 0.31 | 0.83 |
| Heroin | | | | | | | -0.01 | -0.24, 0.22 | 0.91 | -0.01 | -0.24, 0.22 | 0.94 |
| Cannabis | | | | | | | -0.10 | -0.33, 0.13 | 0.37 | -0.11 | -0.34, 0.11 | 0.33 |
| IV drug use | | | | | | | -0.06 | -0.34, 0.22 | 0.66 | -0.01 | -0.29, 0.26 | 0.92 |

| | | | | | | | | | | | | |
|---|---|---|---|---|---|---|---|---|---|---|---|---|
| HIV positivity | | | | | | | | | | -0.14 | -0.46, 0.17 | 0.37 |
| Traumatic brain injury | | | | | | | | | | -0.36 | -0.57, -0.14 | 0.0014 |
| *NOS = not otherwise specified | | | | | | | | | | | | |

**Supplementary Table S7. Full results of factors associated with whole-brain fractional anisotropy (FA) in the precariously housed sample.**

| Average whole-brain fractional anisotropy (FA) | Block 1: Basic demographics | | | Block 2: Mental health diagnoses | | | Block 3: Substance use | | | Block 4: HIV and traumatic brain injury | | |
|---|---|---|---|---|---|---|---|---|---|---|---|---|
| | Beta | 95% CI | *p*-value | Beta | 95% CI | *p*-value | Beta | 95% CI | *p*-value | Beta | 95% CI | *p*-value |
| Age | 0.48 | -0.30, 1.27 | 0.23 | 0.46 | -0.35, 1.27 | 0.27 | 0.31 | -0.55, 1.16 | 0.48 | 0.34 | -0.52, 1.19 | 0.44 |
| Age$^2$ | -1.11 | -1.90, -0.33 | 0.0056 | -1.11 | -1.92, -0.30 | 0.0075 | -0.96 | -1.80, -0.13 | 0.024 | -0.98 | -1.82, -0.14 | 0.022 |
| Sex | | | | | | | | | | | | |
| Female | — | — | | — | — | | — | — | | — | — | |
| Male | 0.20 | -0.07, 0.47 | 0.15 | 0.23 | -0.05, 0.51 | 0.11 | 0.16 | -0.12, 0.45 | 0.25 | 0.16 | -0.12, 0.44 | 0.27 |
| Schizophrenia | | | | -0.24 | -0.63, 0.16 | 0.24 | -0.3 | -0.70, 0.09 | 0.13 | -0.31 | -0.70, 0.08 | 0.12 |
| Schizoaffective | | | | 0.16 | -0.21, 0.53 | 0.4 | 0.13 | -0.24, 0.50 | 0.5 | 0.1 | -0.27, 0.48 | 0.58 |
| Bipolar I or Bipolar NOS | | | | -0.20 | -0.62, 0.23 | 0.36 | -0.17 | -0.58, 0.25 | 0.43 | -0.16 | -0.58, 0.26 | 0.46 |
| Bipolar II | | | | 0.06 | -0.49, 0.61 | 0.83 | 0.10 | -0.45, 0.65 | 0.72 | 0.08 | -0.48, 0.63 | 0.78 |
| Major depressive disorder or depression NOS | | | | -0.07 | -0.4, 0.26 | 0.67 | 0.02 | -0.31, 0.36 | 0.9 | 0.04 | -0.30, 0.37 | 0.84 |
| Psychotic disorder NOS | | | | -0.05 | -0.43, 0.32 | 0.78 | -0.01 | -0.38, 0.36 | 0.95 | 0.01 | -0.36, 0.38 | 0.96 |
| Alcohol | | | | | | | -0.22 | -0.51, 0.07 | 0.13 | -0.23 | -0.52, 0.05 | 0.11 |
| Stimulant | | | | | | | 0.33 | 0.02, 0.64 | 0.035 | 0.33 | 0.02, 0.64 | 0.038 |

| | | | | | | | | | | | | |
|---|---|---|---|---|---|---|---|---|---|---|---|---|
| Heroin | | | | | | | -0.28 | -0.53, -0.03 | 0.027 | -0.29 | -0.55, -0.04 | 0.021 |
| Cannabis | | | | | | | 0.18 | -0.06, 0.43 | 0.15 | 0.19 | -0.06, 0.44 | 0.14 |
| IV drug use | | | | | | | -0.01 | -0.32, 0.29 | 0.93 | 0.01 | -0.29, 0.32 | 0.94 |
| HIV positivity | | | | | | | | | | -0.20 | -0.56, 0.16 | 0.27 |
| Traumatic brain injury | | | | | | | | | | -0.09 | -0.33, 0.15 | 0.46 |

*NOS = not otherwise specified
